# A new model of unreported COVID-19 cases outperforms three known epidemic-growth models in describing data from Cuba and Spain

**DOI:** 10.1101/2021.06.29.21259707

**Authors:** Erick E. Ramirez-Torres, Antonio R. Selva Castañeda, Luis Randez, Luis E. Valdés García, Luis E. Bergues Cabrales, Scott A. Sisson, Juan I. Montijano

## Abstract

Estimating the unreported cases of Covid-19 in a region/country is a complicated problem. We propose a new mathematical model that, combined with a deterministic model of the total growth of cases, describes the time evolution of the unreported cases for each reported Covid-19 case. The new model considers the growth of unreported cases in plateau periods and the decrease towards the end of an epidemic wave. We combined the new model with a Gompertz-growth model, a generalized logistic model, and a susceptible-infectious-removed (SIR) model; and fitted them via Bayesian methods to data from Cuba and Spain. The combined-model fits yielded better Bayesian-Information-Criterion values than the Gompertz, logistic, and SIR models alone. This suggests the new model can achieve improved descriptions of the evolution of a Covid-19 epidemic wave. The new model is also able to provide reliable predictions of the epidemic evolution in a short period of time. We include in the paper the steps that researchers should take to use the new model for predictions with other data.

## 1 Introduction

Estimating the unreported novel coronavirus (SARS-CoV-2) infections is a major challenge for researchers around the world, and it is important in order to give advice to decision-makers. Several studies show that the reporting rate of SARS-CoV-2 infections is variable over time, for reasons that cannot be attributed solely to the transmission dynamics of the Covid-19 (the disease caused by SARS-CoV-2) epidemic [1–5]. This variability can be influenced by the availability of medical supplies, changes in the policies for conducting and reporting tests, hospital capacity, eligibility criteria for people who are tested, among other factors. The data of the Covid-19 epidemic in Wuhan, China, showed an initial peak of 41 infected in the month of December, 2019. Subsequently, for unknown reasons, Wuhan remained without reporting new cases from January 1 to January 15, 2020. This was followed by a huge explosion of cases that has been widely studied and reported [2, 6, 7]. In the province of Santiago de Cuba, Cuba, there was a first wave of cases that had certain similarities with Wuhan, but with a much lower transmissibility [8]. The first infected case was reported on March 27, 2020, and the second case in the next six days. This was followed by several days in which reported infections increased with some regularity. Santiago de Cuba had another period without reporting new cases from April 7 to April 10, 2020, followed by the days of greatest transmissibility in the province until reaching a total of 49 cases in the first wave. This period of several days without registering new cases reported both in Wuhan and in Santiago de Cuba, is not difficult to find in other regions of the world, and they cause plateau areas in the curves of cumulative cases.

From mathematical point of view, uncertainty in the number of undetected infections can lead to misleading inferences and predictions. To be of practical use, models must be able to deal in some way with the difference between reported and actual cases, and with the possible growth of undetected cases during the plateau periods of those detected. An illustrative example of the difficulty of addressing this problem mathematically is that it is impossible with an SI (susceptible-infectious) model to estimate the ratio between reported and unreported cases due to the low identifiability of the model, which provides the same fit to the data for different ratio values [9].

Approaches based on directly fitting a compartmentalized model to the reported data [10–12], or considering the unreported infected one compartment of the model [13,14], cannot capture the recording-no-new-cases periods described in Wuhan and in Santiago de Cuba data, because in these models the derivatives are zeros only in the endemic and disease-free equilibria. An interesting idea to estimate the unreported cases is fitting a susceptible-exposed-infectious-recovered-dead (SEIRD) model directly to numbers of deceased patients, as these are data that provide greater certainty [15]. However, this approach is not useful for predictions in early stages, when there have been very few or no deaths. The exponential-growth fitting used in [2] to evaluate the under-reporting is not feasible when a few plateau days have elapsed while an epidemic wave is fully transmissible.

The aim of this paper is to present a new mathematical model of unreported COVID-19 cases that captures the irregularity of the data when the number of infections is in full growth. This model considers that the total number of cases, both reported and unreported, follows a deterministic law, and it can be combined with known epidemic-growth models. The model also gives reasonable predictions of the epidemic evolution in short periods of time. The paper is structured as follows: the methods section describes the data, the new model, the tested deterministic models, the fitting method, and the criteria for evaluating the results. It is followed by the results section where the fits and quantitative values of the evaluation are shown. The discussion delves into the findings, advantages and limitations of the model. We finish with our conclusions.

## 2 Methods

### 2.1 Data

We used five data sets to examine the performance of the models: the first wave of Covid-19 in Santiago de Cuba (March 20^th^ to May 4^th^, 2020), the first wave of Covid-19 in Cuba (March 11^th^ to July 8^th^, 2020), the second wave of Covid-19 in Santiago de Cuba (October 27^th^, 2020 to January 1^st^, 2021), the second wave of Covid-19 in Cuba (August 7^th^ to October 6^th^, 2020), and the first wave of Covid-19 in Spain (February 20^th^ to May 17^th^, 2020). Cuban data were supplied by the Ministry of Public Health (https://covid19cubadata.github.io), and data from Spain were issued by the Ministry of Health, Consumption and Social Welfare (https://cnecovid.isciii.es/covid19).

### 2.2 Models

We argue that the number of unreported Covid-19 cases for each reported case (*U* (*n*)) depends on the difference between the accumulated cases reported until the current day (*CR*(*n*)) and those reported the previous day (*CR*(*n* − 1)), as well as the increase in active cases (*I*(*n*) − *I*(*n* − 1)). We propose the model

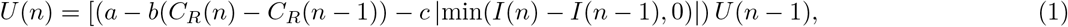

where *C*_*R*_(*n*) is the reported number of cumulative cases at day *n, I*(*n*) is the number of active cases at day *n, a* ≥ 1 is the unreported-growth rate (this parameter captures the rate of the increment of cases when no new case is detected), *b* ≥ 0 is a coefficient that is related to the effectiveness in detecting new cases, and *c*, if positive, captures a possible decrease in the fraction of unreported cases towards the end of an epidemic wave. If the decrease in active cases is due more to under-detection than to the influence of the end of the wave, *c* takes a negative value.

This model accounts for the under-reporting that is likely in the early stages of the epidemic [2]. In those early stages, if a plateau occurs, the difference *C*_*R*_(*n*) −*C*_*R*_(*n* −1) is zero or close to zero, and the value of *U* (*n*) grows proportional to *a*. When an epidemic wave nears its end, the number of active cases decreases, but it is to be hoped that the authorities have improved the mechanisms for detecting new infections. For this reason the model contains the term with the coefficient *c*, which exerts influence in decreasing sections of the active cases that occur at the end of the wave. Equation (1) can complement a deterministic model of infections, including detected and undetected cases. To describe the dynamics of the total cumulative-case number, we use three phenomenological models that have been previously applied in the forecast of infectious disease outbreaks.

The first phenomenological model is an extension of the Richards growth model [16], given by

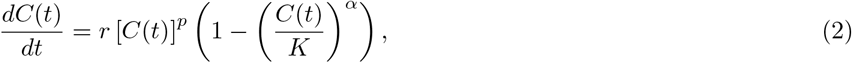

where *t* is the time, *r* ≥ 0 is the growth rate, *p* (0 ≤ *p* ≤ 1) represents the deceleration of growth, *K >* 0 is the size of the epidemic, and *α*(0 ≤ *α* ≤ 1) is a parameter that captures the extent of deviation from the S-shaped dynamics of the classical logistic growth model [17]. This model is known as generalized Richard model (GRM), and it has been successfully used to make post-peak predictions of several infectious disease epidemics [18–20].

The second phenomenological model is the Gompertz curve, which has been used to describe several biological growths [21], including the Covid-19 epidemic [22, 23]. The parametrisation of Gompertz growth that we use is

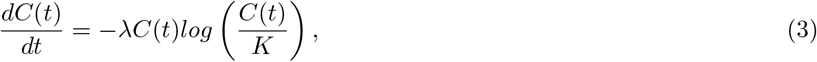

where *λ >* 0 is the growth rate. Compared with GRM, the Gompertz curve changes faster early but approaches the asymptote more slowly. The third model is a conventional SIR [24] given by

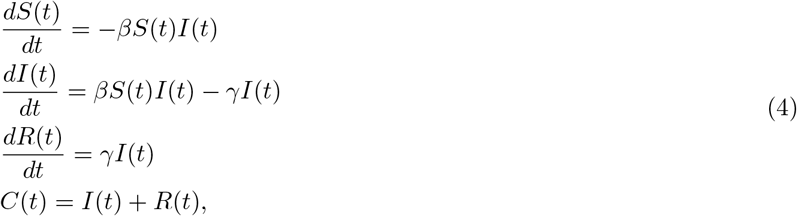

where *S*(*t*) is the number of susceptible individuals at time *t*; *I*(*t*) is the number of infected individuals at time *t*; *R*(*t*) is the number of removed (both recovered and dead) individuals at time *t*; and *β* and *γ* are the transmission rate and rate of removal, respectively.

If the numerical solution of the phenomenological model (obtained from equation 2, 3 or 4) at day *n* is denoted as *C*(*n*), the reported number of cumulative cases *C*_*R*_(*n*) and the unreported ratio *U* (*n*) are related by

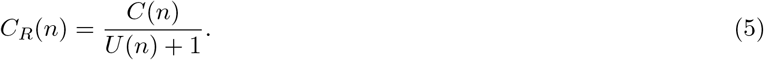

In the following we refer to the combination of equations 1, 2 and 5 as GRM+U; the combination of equations 1, 3 and 5 as Gompertz+U; and the combination of equations 1, 4 and 5 as SIR+U. For comparison, the GRM (equation 2), Gompertz (equation 3) and SIR (equation 4) models were fitted to the data.

### 2.3 Model fitting and assessment

We explored several fits using a Student’s t distribution of the error in case observation [25] and the best results were obtained with high values of the degree-of-freedom parameter (*v*). For high values of *v*, the Student’s t distribution resembles the Gaussian distribution. For this reason we prefer to use a normal error, which in turn has other advantages [26–28], and is given by

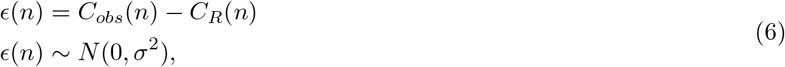

where *C*_*obs*_(*n*) is the observed value of cumulative cases at day *n*. Although *C*_*R*_(*n*) is a discrete variable, *ϵ* (*n*) is continuous, and the assumption that it has a normal distribution facilitates the likelihood estimation. On the other hand, we assume a constant variance instead of a variance proportional to the mean as in [25]. We consider that the observation noise through random subsampling is not necessarily greater in the final days of an epidemic wave than at the beginning of it, but rather has a more complicated variability that depends on the quality of the epidemiological investigation. This variability is already considered in the rationale of the new proposed model (equation 1).

We infer the parameters with the Bayesian approach. To represent the little information available on the parameters, we introduced improper uniform priors in all models (Tables 1-15). In the SIR models, it was necessary to introduce proper informative priors for parameters *β* and *γ* (Tables 3, 6, 9, 12 and 15), because it was practically impossible to obtain reasonable posteriors using uninformative priors. This is consistent with the SIR model being practically non-identifiable when only cumulative-case data are used [29]. The means of the prior distributions of parameters *β* and *γ* were determined empirically.

**Table 1:** Summary table of gompertz-based models fitted to the data of the first epidemic wave in Santiago de Cuba.

| Model | Parameter | Prior or fixed value | Mean of posterior (95% credible interval) | BIC |
| --- | --- | --- | --- | --- |
| *Gompertz* | $\lambda$ | $U(0, \infty)$ | 0.3137 (0.1624, 0.5103) | 175 |
| | $K$ | 49 | - | |
| | $C(0)$ | 1 | - | |
| | $\sigma$ | $U(0, \infty)$ | 2.4824 (1.2428, 4.2252) | |
| *Gompertz + U* | $\lambda$ | $U(0, \infty)$ | 0.3137 (0.1624, 0.5103) | 101 |
| | $K$ | $U(49, \infty)$ | 61.4700 (49.2515, 76.4326) | |
| | $C(0)$ | $U(1, 20)$ | 3.7550 (1.0026, 8.8356) | |
| | $a$ | $U(1, \infty)$ | 1.0381 (1.0025, 1.0726) | |
| | $b$ | $U(0, \infty)$ | $8.5422 \times 10^{-2}$ ($5.7316 \times 10^{-2}$, $1.1333 \times 10^{-1}$) | |
| | $c$ | $U(-\infty, \infty)$ | $1.6394 \times 10^{-2}$ ($-4.8427 \times 10^{-3}$, $5.6142 \times 10^{-2}$) | |
| | $U(0)$ | $U(0, 19)$ | 7.4336 (4.5499, 10.5675) | |
| | $\sigma$ | $U(0, \infty)$ | 0.9475 (0.4886, 1.6215) | |

**Table 2:** Summary table of GRM-based models fitted to the data of the first epidemic wave in Santiago de Cuba.

| Model | Parameter | Prior or fixed value | Mean of posterior (95% credible interval) | BIC |
| --- | --- | --- | --- | --- |
| *GRM* | $r$ | $U(0, \infty)$ | 0.6488 (0.2347, 1.3566) | 183 |
| | $\alpha$ | $U(0, 1)$ | 0.5124 (0.1283, 0.9623) | |
| | $p$ | $U(0, 1)$ | 0.7847 (0.4745, 0.9998) | |
| | $K$ | 49 | - | |
| | $C(0)$ | 1 | - | |
| | $\sigma$ | $U(0, \infty)$ | 2.8360 (1.2417, 5.3519) | |
| *GRM + U* | $r$ | $U(0, \infty)$ | 3.6206 (1.1376, 8.4284) | 90 |
| | $\alpha$ | $U(0, 1)$ | $4.8178 \times 10^{-1}$ ($5.5569 \times 10^{-2}$, $9.5179 \times 10^{-1}$) | |
| | $p$ | $U(0, 1)$ | $2.1982 \times 10^{-1}$ ($2.5707 \times 10^{-3}$, $5.9589 \times 10^{-1}$) | |
| | $K$ | $U(49, \infty)$ | 72.7841 (52.5528, 89.7081) | |
| | $C(0)$ | $U(1, 20)$ | 5.5029 (1.0013, 15.7326) | |
| | $a$ | $U(1, \infty)$ | 1.1158 (1.0018, 1.2064) | |
| | $b$ | $U(0, \infty)$ | $1.4225 \times 10^{-1}$ ($7.8214 \times 10^{-2}$, $1.9219 \times 10^{-1}$) | |
| | $c$ | $U(-\infty, \infty)$ | $7.8667 \times 10^{-7}$ ($-2.0816 \times 10^{-5}$, $3.4945 \times 10^{-5}$) | |
| | $U(0)$ | $U(0, 19)$ | 3.7705 (0.9020, 10.0773) | |
| | $\sigma$ | $U(0, \infty)$ | 1.1383 (0.5180, 2.3713) | |

**Table 3:** Summary table of SIR-based models fitted to the data of the first epidemic wave in Santiago de Cuba.

| Model | Parameter | Prior or fixed value | Mean of posterior (95% credible interval) | BIC |
| --- | --- | --- | --- | --- |
| *SIR* | $\beta$ | $LogNormal(\log(0.22), 0.5)$ | 1.7064 ($1.0304 \times 10^{-2}$, 5.0997) | 254 |
| | $\gamma$ | $LogNormal(\log(0.12), 0.5)$ | 2.8674 ($8.5869 \times 10^{-3}$, 12.4458) | |
| | $I(0)$ | 1 | - | |
| | $R(0)$ | 0 | - | |
| | $\sigma$ | $U(0, \infty)$ | 21.0016(1.7813, 66.8076) | |
| *SIR + U* | $\beta$ | $U(0, \infty)$ | 0.9744 (0.3893, 1.7671) | 111 |
| | $\gamma$ | $U(0, \infty)$ | 0.8932 (0.3543, 1.6087) | |
| | $a$ | $U(1, \infty)$ | 1.0977 (1.0000, 1.3109) | |
| | $b$ | $U(0, \infty)$ | 0.2326 (0.1223, 0.4399) | |
| | $c$ | $U(-\infty, \infty)$ | -2.1319 (-15.1956, 2.0142) | |
| | $U(0)$ | $U(0, 19)$ | 4.1938 (0.1301, 10.5409) | |
| | $I(0)$ | $U(1, 20)$ | 2.2347 (1.0028, 3.6856) | |
| | $R(0)$ | 0 | - | |
| | $\sigma$ | $U(0, \infty)$ | 1.8779 (0.6149, 4.3839) | |

**Table 4:**
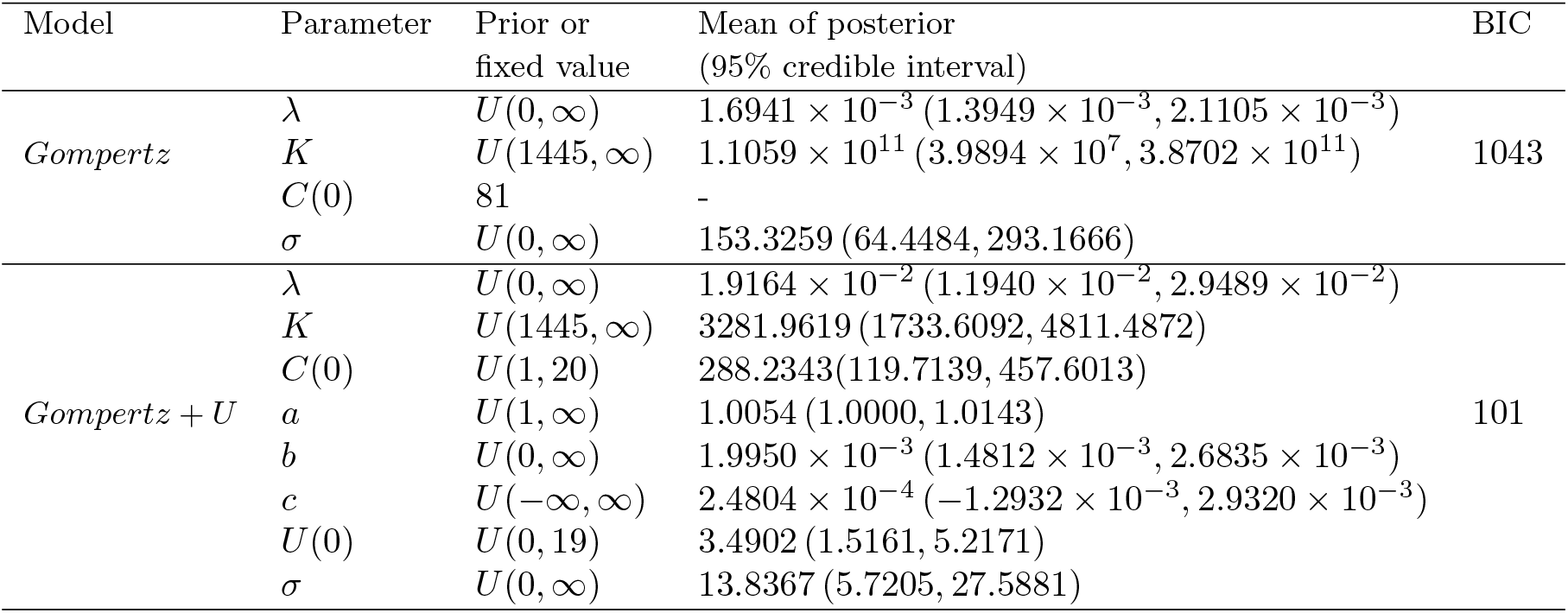
Summary table of gompertz-based models fitted to the data of the second epidemic wave in Santiago de Cuba.

| Model | Parameter | Prior or fixed value | Mean of posterior (95% credible interval) | BIC |
| --- | --- | --- | --- | --- |
| *Gompertz* | $\lambda$ | $U(0, \infty)$ | $1.6941 \times 10^{-3}$ ($1.3949 \times 10^{-3}$, $2.1105 \times 10^{-3}$) | 1043 |
| | $K$ | $U(1445, \infty)$ | $1.1059 \times 10^{11}$ ($3.9894 \times 10^7$, $3.8702 \times 10^{11}$) | |
| | $C(0)$ | 81 | - | |
| | $\sigma$ | $U(0, \infty)$ | 153.3259 (64.4484, 293.1666) | |
| *Gompertz + U* | $\lambda$ | $U(0, \infty)$ | $1.9164 \times 10^{-2}$ ($1.1940 \times 10^{-2}$, $2.9489 \times 10^{-2}$) | 101 |
| | $K$ | $U(1445, \infty)$ | 3281.9619 (1733.6092, 4811.4872) | |
| | $C(0)$ | $U(1, 20)$ | 288.2343 (119.7139, 457.6013) | |
| | $a$ | $U(1, \infty)$ | 1.0054 (1.0000, 1.0143) | |
| | $b$ | $U(0, \infty)$ | $1.9950 \times 10^{-3}$ ($1.4812 \times 10^{-3}$, $2.6835 \times 10^{-3}$) | |
| | $c$ | $U(-\infty, \infty)$ | $2.4804 \times 10^{-4}$ ($-1.2932 \times 10^{-3}$, $2.9320 \times 10^{-3}$) | |
| | $U(0)$ | $U(0, 19)$ | 3.4902 (1.5161, 5.2171) | |
| | $\sigma$ | $U(0, \infty)$ | 13.8367 (5.7205, 27.5881) | |

**Table 5:** Summary table of GRM-based models fitted to the data of the second epidemic wave in Santiago de Cuba.

| Model | Parameter | Prior or fixed value | Mean of posterior (95% credible interval) | BIC |
| --- | --- | --- | --- | --- |
| *GRM* | $r$ | $U(0, \infty)$ | 4.2878 (0.1096, 10.2579) | 1089 |
| | $\alpha$ | $U(0, 1)$ | $4.7975 \times 10^{-1}$ ($3.3639 \times 10^{-3}$, $9.4257 \times 10^{-1}$) | |
| | $p$ | $U(0, 1)$ | $2.1520 \times 10^{-1}$ ($1.6234 \times 10^{-4}$, $6.7513 \times 10^{-1}$) | |
| | $K$ | $U(1445, \infty)$ | $5.9624 \times 10^{15}$ ($1.7468 \times 10^{11}$, $5.2337 \times 10^{16}$) | |
| | $C(0)$ | 81 | - | |
| | $\sigma$ | $U(0, \infty)$ | 13.8367 (5.7205, 27.5881) | |
| *GRM + U* | $r$ | $U(0, \infty)$ | $1.4868 \times 10^{-1}$ ($3.3457 \times 10^{-2}$, $3.1653 \times 10^{-1}$) | 608 |
| | $\alpha$ | $U(0, 1)$ | 0.6860 (0.3827, 0.9998) | |
| | $p$ | $U(0, 1)$ | 0.8414 (0.6861, 0.9998) | |
| | $K$ | $U(1445, \infty)$ | 3744.6757 (2114.7993, 6801.9879) | |
| | $C(0)$ | $U(81, 1701)$ | 291.8661 (106.7113, 453.6048) | |
| | $a$ | $U(1, \infty)$ | 1.0063 (1.0000, 1.0183) | |
| | $b$ | $U(0, \infty)$ | $1.9197 \times 10^{-3}$ ($1.4441 \times 10^{-3}$, $2.3451 \times 10^{-3}$) | |
| | $c$ | $U(-\infty, \infty)$ | $-3.5555 \times 10^{-7}$ ($-5.3732 \times 10^{-5}$, $6.7889 \times 10^{-5}$) | |
| | $U(0)$ | $U(0, 20)$ | 3.2111 (0.9658, 4.9073) | |
| | $\sigma$ | $U(0, \infty)$ | 12.1945 (5.5759, 22.2659) | |

**Table 6:** Summary table of SIR-based models fitted to the data of the second epidemic wave in Santiago de Cuba.

| Model | Parameter | Prior or fixed value | Mean of posterior (95% credible interval) | BIC |
| --- | --- | --- | --- | --- |
| *SIR* | $\beta$ | $LogNormal(\log(5), 0.5)$ | 2.0762 (0.3836, 4.6689) | 943 |
| | $\gamma$ | $LogNormal(\log(4.5), 0.5)$ | 1.8243 (0.2719, 4.1686) | |
| | $I(0)$ | 1 | - | |
| | $R(0)$ | 80 | - | |
| | $\sigma$ | $U(0, \infty)$ | 102.0311 (33.3842, 207.0680) | |
| *SIR + U* | $\beta$ | $U(0, \infty)$ | 19.1045 (0.5972, 53.8317) | 815 |
| | $\gamma$ | $U(0, \infty)$ | 18.9268 (0.5821, 53.1176) | |
| | $a$ | $U(1, \infty)$ | 1.0127 (1.0000, 1.0365) | |
| | $b$ | $U(0, \infty)$ | $3.1095 \times 10^{-3}$ ($2.2763 \times 10^{-3}$, $4.1646 \times 10^{-3}$) | |
| | $c$ | $U(-\infty, \infty)$ | $9.2737 \times 10^{-3}$ ($1.7625 \times 10^{-3}$, $1.8085 \times 10^{-2}$) | |
| | $U(0)$ | $U(0, 20)$ | 10.3220 (5.1159, 14.7504) | |
| | $I(0)$ | $U(1, 1701)$ | 120.4135 (97.4859, 158.0629) | |
| | $R(0)$ | 80 | - | |
| | $\sigma$ | $U(0, \infty)$ | 45.3306 (23.4194, 66.7243) | |

**Table 7:** Summary table of Gompertz-based models fitted to the data of the first epidemic wave in Cuba.

| Model | Parameter | Prior or fixed value | Mean of posterior (95% credible interval) | BIC |
| --- | --- | --- | --- | --- |
| *Gompertz* | $\lambda$ | $U(0, \infty)$ | $5.1734 \times 10^{-2}$ ($4.8491 \times 10^{-2}$, $5.4937 \times 10^{-2}$) | 1349 |
| | $K$ | $U(2403, \infty)$ | 2431.2304 (2403.0064, 2488.7812) | |
| | $C(0)$ | 3 | - | |
| | $\sigma$ | $U(0, \infty)$ | 89.5801 (45.6778, 147.3408) | |
| *Gompertz + U* | $\lambda$ | $U(0, \infty)$ | $9.4596 \times 10^{-2}$ ($4.3327 \times 10^{-2}$, $1.6541 \times 10^{-1}$) | 1006 |
| | $K$ | $U(2403, \infty)$ | 6096.4097 (2646.4303, 10642.0817) | |
| | $C(0)$ | $U(3, 63)$ | 43.6962 (30.2954, 55.5618) | |
| | $a$ | $U(1, \infty)$ | 1.0017 (1.0000, 1.0059) | |
| | $b$ | $U(0, \infty)$ | $9.6064 \times 10^{-4}$ ($4.1484 \times 10^{-4}$, $1.7985 \times 10^{-3}$) | |
| | $c$ | $U(-\infty, \infty)$ | $-1.5727 \times 10^{-4}$ ($-5.8128 \times 10^{-4}$, $1.6325 \times 10^{-4}$) | |
| | $U(0)$ | $U(0, 20)$ | 9.7985 (2.9344, 16.4708) | |
| | $\sigma$ | $U(0, \infty)$ | 40.3523 (10.9110, 88.0362) | |

**Table 8:** Summary table of GRM-based models fitted to the data of the first epidemic wave in Cuba.

| Model | Parameter | Prior or fixed value | Mean of posterior (95% credible interval) | BIC |
| --- | --- | --- | --- | --- |
| *GRM* | $r$ | $U(0, \infty)$ | 1.5817 (0.3961, 3.9349) | 1327 |
| | $\alpha$ | $U(0, 1)$ | $4.4193 \times 10^{-1}$ ($4.6058 \times 10^{-2}$, $9.3341 \times 10^{-1}$) | |
| | $p$ | $U(0, 1)$ | 0.6871 (0.4109, 0.9201) | |
| | $K$ | $U(2403, \infty)$ | 2557.8669 (2403.2109, 2939.1628) | |
| | $C(0)$ | 3 | - | |
| | $\sigma$ | $U(0, \infty)$ | 102.4544 (43.3811, 193.0814) | |
| *GRM + U* | $r$ | $U(0, \infty)$ | 15.6206 (2.6769, 31.1859) | 1321 |
| | $\alpha$ | $U(0, 1)$ | $4.7957 \times 10^{-1}$ ($7.2105 \times 10^{-2}$, $8.8309 \times 10^{-1}$) | |
| | $p$ | $U(0, 1)$ | $2.4900 \times 10^{-1}$ ($6.0741 \times 10^{-2}$, $4.9375 \times 10^{-1}$) | |
| | $K$ | $U(2403, \infty)$ | 3976.3169 (3942.6274, 4000.2629) | |
| | $C(0)$ | $U(3, 63)$ | 4.8442 (3.0029, 8.2446) | |
| | $a$ | $U(1, \infty)$ | 1.0632 (1.0002, 1.1443) | |
| | $b$ | $U(0, \infty)$ | $5.8882 \times 10^{-3}$ ($2.2049 \times 10^{-3}$, $1.0048 \times 10^{-2}$) | |
| | $c$ | $U(-\infty, \infty)$ | $-5.1432 \times 10^{-3}$ ($-1.1118 \times 10^{-2}$, $-1.9149 \times 10^{-4}$) | |
| | $U(0)$ | $U(0, 20)$ | 4.2543 (0.6039, 8.8050) | |
| | $\sigma$ | $U(0, \infty)$ | 93.9434 (81.5936, 103.3677) | |

**Table 9:** Summary table of SIR-based models fitted to the data of the first epidemic wave in Cuba.

| Model | Parameter | Prior or fixed value | Mean of posterior (95% credible interval) | BIC |
| --- | --- | --- | --- | --- |
| *SIR* | $\beta$ | $LogNormal(\log(11), 0.5)$ | 13.2938 (7.2963, 20.4458) | 1632 |
| | $\gamma$ | $LogNormal(\log(10), 0.5)$ | 11.9727 (6.5665, 18.4243) | |
| | $I(0)$ | 3 | - | |
| | $R(0)$ | 0 | - | |
| | $\sigma$ | $U(0, \infty)$ | 285.3559 (133.3871, 529.4842) | |
| *SIR + U* | $\beta$ | $U(0, \infty)$ | 2.7116 (0.9531, 4.9841) | 1255 |
| | $\gamma$ | $U(0, \infty)$ | 2.4553 (0.8685, 4.5003) | |
| | $a$ | $U(1, \infty)$ | 1.0566 (1.0001, 1.2201) | |
| | $b$ | $U(0, \infty)$ | $5.0813 \times 10^{-3}$ ($2.2823 \times 10^{-3}$, $9.8844 \times 10^{-3}$) | |
| | $c$ | $U(-\infty, \infty)$ | $3.5628 \times 10^{-2}$ ($-3.8565 \times 10^{-3}$, $8.0601 \times 10^{-2}$) | |
| | $U(0)$ | $U(0, 20)$ | 6.2663 ($6.1497 \times 10^{-2}$, 14.8329) | |
| | $I(0)$ | $U(3, 63)$ | 24.3205 (9.1143, 42.6814) | |
| | $R(0)$ | 0 | - | |
| | $\sigma$ | $U(0, \infty)$ | 80.6344 (56.8924, 108.7357) | |

**Table 10:** Summary table of Gompertz-based models fitted to the data of the second epidemic wave in Cuba.

| Model | Parameter | Prior or fixed value | Mean of posterior (95% credible interval) | BIC |
| --- | --- | --- | --- | --- |
| *Gompertz* | $\lambda$ | $U(0, \infty)$ | $9.9352 \times 10^{-3}$ ($5.2862 \times 10^{-3}$, $1.3703 \times 10^{-2}$) | 599 |
| | $K$ | $U(5898, \infty)$ | $1.6463 \times 10^4$ ($8.3706 \times 10^3$, $3.1391 \times 10^4$) | |
| | $C(0)$ | 2953 | - | |
| | $\sigma$ | $U(0, \infty)$ | 65.2692 (18.5353, 145.5541) | |
| *Gompertz + U* | $\lambda$ | $U(0, \infty)$ | $9.9554 \times 10^{-3}$ ($1.3711 \times 10^{-3}$, $1.8247 \times 10^{-2}$) | 406 |
| | $K$ | $U(5898, \infty)$ | $2.4198 \times 10^4$ ($7.9431 \times 10^3$, $4.6837 \times 10^4$) | |
| | $C(0)$ | $U(2953, 62013)$ | $7.9112 \times 10^3$ ($3.4848 \times 10^3$, $1.1504 \times 10^4$) | |
| | $a$ | $U(1, \infty)$ | 1.0228 (1.0000, 1.0797) | |
| | $b$ | $U(0, \infty)$ | $7.8182 \times 10^{-4}$ ($2.0946 \times 10^{-4}$, $2.2427 \times 10^{-3}$) | |
| | $c$ | $U(-\infty, \infty)$ | $-4.8757 \times 10^{-5}$ ($-8.6359 \times 10^{-4}$, $1.0701 \times 10^{-3}$) | |
| | $U(0)$ | $U(0, 20)$ | 1.6620 (0.1584, 2.8782) | |
| | $\sigma$ | $U(0, \infty)$ | 32.7781 (5.1966, 68.3806) | |

**Table 11:** Summary table of GRM-based models fitted to the data of the second epidemic wave in Cuba.

| Model | Parameter | Prior or fixed value | Mean of posterior (95% credible interval) | BIC |
| --- | --- | --- | --- | --- |
| *GRM* | $r$ | $U(0, \infty)$ | 1.3800 ($4.6876 \times 10^{-2}$, 4.2984) | 607 |
| | $\alpha$ | $U(0, 1)$ | 0.5771 (0.1191, 0.9909) | |
| | $p$ | $U(0, 1)$ | 0.5691 (0.3103, 0.8126) | |
| | $K$ | $U(5898, \infty)$ | $1.6412 \times 10^4$ ($1.5709 \times 10^4$, $1.8674 \times 10^4$) | |
| | $C(0)$ | 2953 | - | |
| | $\sigma$ | $U(0, \infty)$ | 48.6199 (22.6212, 86.7442) | |
| *GRM + U* | $r$ | $U(0, \infty)$ | 9.2802 ($8.1602 \times 10^{-2}$, 34.0746) | 498 |
| | $\alpha$ | $U(0, 1)$ | $4.4517 \times 10^{-1}$ ($1.4434 \times 10^{-4}$, $9.2898 \times 10^{-1}$) | |
| | $p$ | $U(0, 1)$ | $3.05874 \times 10^{-1}$ ($2.5280 \times 10^{-3}$, $6.7356 \times 10^{-1}$) | |
| | $K$ | $U(5898, \infty)$ | $2.0019 \times 10^4$ ($1.9828 \times 10^4$, $2.0224 \times 10^4$) | |
| | $C(0)$ | $U(2953, 62013)$ | 5917.2175 (5905.2430, 5957.8293) | |
| | $a$ | $U(1, \infty)$ | 1.0541 (1.0000, 1.1884) | |
| | $b$ | $U(0, \infty)$ | $1.4962 \times 10^{-3}$ ($9.3454 \times 10^{-6}$, $3.9971 \times 10^{-3}$) | |
| | $c$ | $U(-\infty, \infty)$ | $4.0979 \times 10^{-4}$ ($-1.9197 \times 10^{-3}$, $3.4022 \times 10^{-3}$) | |
| | $U(0)$ | $U(0, 20)$ | 1.0386 (0.7786, 1.3217) | |
| | $\sigma$ | $U(0, \infty)$ | 101.4516 (16.8961, 240.0584) | |

**Table 12:**
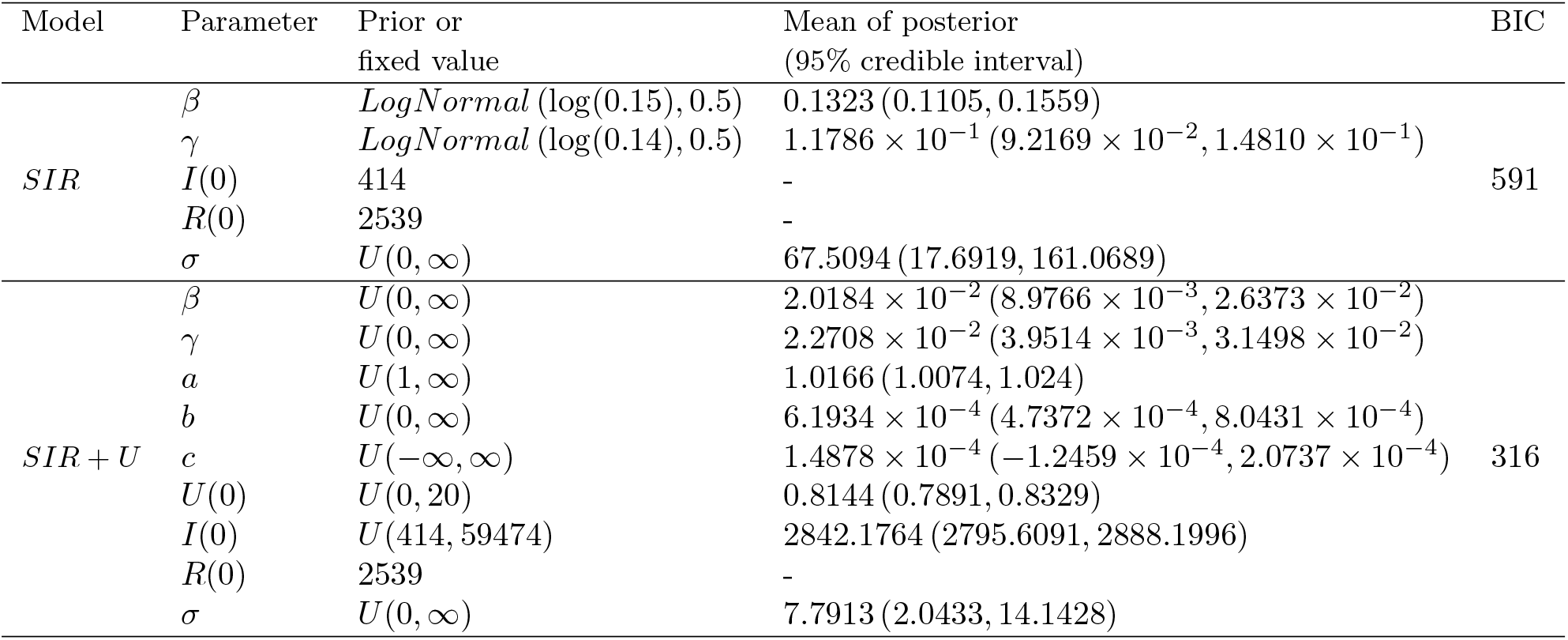
Summary table of SIR-based models fitted to the data of the second epidemic wave in Cuba.

**Table 13:** Summary table of Gompertz-based models fitted to the data of the first epidemic wave in Spain.

| Model | Parameter | Prior or fixed value | Mean of posterior (95% credible interval) | BIC |
| --- | --- | --- | --- | --- |
| *Gompertz* | $\lambda$ | $U(0, \infty)$ | $7.2171 \times 10^{-2}$ ($3.7032 \times 10^{-5}$, $1.7174 \times 10^{-1}$) | 1800 |
| | $K$ | $U(2.3 \times 10^5, \infty)$ | $1.5268 \times 10^6$ ($2.3164 \times 10^5$, $8.4524 \times 10^6$) | |
| | $C(0)$ | 3 | - | |
| | $\sigma$ | $U(0, \infty)$ | $1.2305 \times 10^5$ ($3.0695 \times 10^3$, $4.8947 \times 10^5$) | |
| *Gompertz + U* | $\lambda$ | $U(0, \infty)$ | $9.3581 \times 10^{-2}$ ($6.4077 \times 10^{-2}$, $1.3402 \times 10^{-1}$) | 1748 |
| | $K$ | $U(2.3 \times 10^5, \infty)$ | $6.6409 \times 10^5$ ($2.3163 \times 10^5$, $3.6676 \times 10^6$) | |
| | $C(0)$ | $U(3, 453)$ | 14.2302 (3.9023, 23.1068) | |
| | $a$ | $U(1, \infty)$ | 1.0260 (1.0001, 1.0663) | |
| | $b$ | $U(0, \infty)$ | $2.2422 \times 10^{-5}$ ($1.1587 \times 10^{-5}$, $4.2329 \times 10^{-5}$) | |
| | $c$ | $U(-\infty, \infty)$ | $9.4064 \times 10^{-6}$ ($-3.1404 \times 10^{-5}$, $4.3426 \times 10^{-5}$) | |
| | $U(0)$ | $U(0, 150)$ | 5.2966 (2.9321, 11.5072) | |
| | $\sigma$ | $U(0, \infty)$ | $9.1109 \times 10^3$ ($5.7753 \times 10^3$, $1.3015 \times 10^4$) | |

**Table 14:** Summary table of GRM-based models fitted to the data of the first epidemic wave in Spain.

| Model | Parameter | Prior or fixed value | Mean of posterior (95% credible interval) | BIC |
| --- | --- | --- | --- | --- |
| *GRM* | $r$ | $U(0, \infty)$ | 2045.7256 (13.1868, 5258.1877) | 2083 |
| | $\alpha$ | $U(0, 1)$ | $3.7832 \times 10^{-1}$ ($7.6067 \times 10^{-5}$, $8.9392 \times 10^{-1}$) | |
| | $p$ | $U(0, 1)$ | $9.8282 \times 10^{-2}$ ($5.2107 \times 10^{-5}$, $2.6587 \times 10^{-1}$) | |
| | $K$ | $U(2.3 \times 10^5, \infty)$ | $1.3737 \times 10^8$ ($2.3422 \times 10^5$, $7.9813 \times 10^8$) | |
| | $C(0)$ | 3 | - | |
| | $\sigma$ | $U(0, \infty)$ | $8.2163 \times 10^4$ ($1.4561 \times 10^4$, $2.1815 \times 10^5$) | |
| *GRM + U* | $r$ | $U(0, \infty)$ | 3.1280 (0.5202, 8.0344) | 1766 |
| | $\alpha$ | $U(0, 1)$ | 0.5919 (0.1316, 0.9999) | |
| | $p$ | $U(0, 1)$ | 0.8488 (0.7288, 0.9595) | |
| | $K$ | $U(2.3 \times 10^5, \infty)$ | $8.2157 \times 10^6$ ($1.8667 \times 10^6$, $2.7209 \times 10^7$) | |
| | $C(0)$ | $U(3, 453)$ | 57.9959 (3.0213, 221.7619) | |
| | $a$ | $U(1, \infty)$ | 1.0366 (1.0000, 1.0843) | |
| | $b$ | $U(0, \infty)$ | $1.1834 \times 10^{-5}$ ($1.1488 \times 10^{-6}$, $2.3482 \times 10^{-5}$) | |
| | $c$ | $U(-\infty, \infty)$ | $1.5851 \times 10^{-5}$ ($-2.7304 \times 10^{-6}$, $3.9027 \times 10^{-5}$) | |
| | $U(0)$ | $U(0, 150)$ | 44.2055 (8.7233, 127.0796) | |
| | $\sigma$ | $U(0, \infty)$ | $7.8890 \times 10^3$ ($6483.2756 \times 10^3$, $1.2120 \times 10^4$) | |

**Table 15:** Summary table of SIR-based models fitted to the data of the first epidemic wave in Spain.

| Model | Parameter | Prior or fixed value | Mean of posterior (95% credible interval) | BIC |
| --- | --- | --- | --- | --- |
| *SIR* | $\beta$ | $LogNormal(\log(28), 0.5)$ | 59.7194 (38.9798, 90.8963) | 1801 |
| | $\gamma$ | $LogNormal(\log(25), 0.5)$ | 53.6142 (34.9657, 81.6334) | |
| | $I(0)$ | 3 | - | |
| | $R(0)$ | 0 | - | |
| | $\sigma$ | $U(0, \infty)$ | $1.9462 \times 10^4$ ($3.8930 \times 10^3$, $4.5512 \times 10^4$) | |
| *SIR + U* | $\beta$ | $U(0, \infty)$ | 45.1051 (6.2945, 90.9143) | 1766 |
| | $\gamma$ | $U(0, \infty)$ | 40.4471 (5.4096, 81.6242) | |
| | $a$ | $U(1, \infty)$ | 1.0383 (1.0000, 1.0964) | |
| | $b$ | $U(0, \infty)$ | $2.2005 \times 10^{-5}$ ($8.2259 \times 10^{-6}$, $3.6131 \times 10^{-5}$) | |
| | $c$ | $U(-\infty, \infty)$ | $3.7172 \times 10^{-5}$ ($-7.562210 - 6$, $1.017810 - 4$) | |
| | $U(0)$ | $U(0, 150)$ | 6.8545 (0.8298, 19.1129) | |
| | $I(0)$ | $U(3, 453)$ | 58.8986 (6.2478, 96.6126) | |
| | $R(0)$ | 0 | - | |
| | $\sigma$ | $U(0, \infty)$ | $6.9022 \times 10^3$ ($6.7437 \times 10^3$, $7.0386 \times 10^3$) | |

As of May 4, 2020, no more cases were registered in Santiago de Cuba for more than 30 days. For this reason, we consider the first wave completely finished on May 4, and we set the value of *K* = 49, both for the Gompertz model and for the GRM model. Also, we set 49 the lower limit of the improper uniform prior of *K* for the Gompertz+U and GRM+U models. Based on the expertise of epidemiologists and on estimates made in previous work [8], we set relatively small upper limits for the priors of C and U (20 and 19, respectively). This upper limit made it possible to reduce the search region of the posterior distributions of parameters and to reduce the risk of practical non-identifiability [29, 30]. Tables 1-3 show details of the priors and the fixed values of the parameters for the fitting of the models to the data of the first wave in Santiago de Cuba.

In the other waves analyzed, a clear ending is not seen, so we empirically cut the data on certain dates to fit the models, and we used the values of cumulative cases on those dates as lower limits of the uniform priors of *K* in all models (1445 for the second wave in Santiago de Cuba, 2403 for the first wave in Cuba, 5898 for the second wave in Cuba, and 2.3 × 10^5^ for the first wave in Spain). The numbers of cumulative cases reported in the initial days of each data set were used either as lower limits of the uniform priors of *C*(0) if the new model (+U) is included, or as fixed values of *C*(0), if the new model is not included. The cumulative cases reported on those initial dates are 81 for the second wave in Santiago de Cuba, 3 for the first wave in Cuba, 2953 for the second wave in Cuba, and 3 for the first wave in Spain. A previous work [31] estimated a value of *U* (*n*) = 6.23 for Cuba for *n* corresponding to the date December 22, 2020. Taking into account that the two waves analyzed in Cuba and the beginning of the second wave in Santiago de Cuba do not exceed the date mentioned above, we decided to establish an empirical upper limit of 20 to the uniform prior of *U* (0) (tables 2, 3 and 4). Due to the large number of cases reported in Spain (the highest value of accumulated cases in the data studied from Spain is more than 30 times higher than the highest number reported in Cuba), we decided to establish an upper limit of 150 to the uniform prior of *U* (0) for models fitted to data from Spain.

In the Bayesian inference carried out, the samples of the posterior distributions of the parameters were obtained using the Delayed-Rejection-Adaptive-Metropolis (DRAM) algorithm, implemented via the Python Package **pymcmcstat** [32], with 5 × 10^5^ samples and burn-in number of 2.5 × 10^5^. We generated 2 × 10^4^ trajectories of the different models by sampling from the inferred posterior distributions of the parameters. From these trajectories we determine a mean of posterior for forecast, and a 95% credible interval for each day. Models were compared using the Bayesian information criterion (BIC) [33], using the maximum a posteriori (MAP) point. The differential-evolution algorithm [34] was used to find the MAP. We chose the BIC because it rewards a good description of the experimental data and penalizes the complexity of the model.

With the data from the second wave in Santiago de Cuba, we extrapolated using the fit of the models to predict the cases reported in the following days. We extrapolated using the function

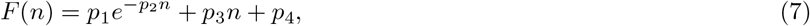

where *n* is the day, *F* is the ratio between the total and reported cumulative cases (*F* (*n*) = *C*(*n*)*/C*_*R*_(*n*) = *U* (*n*) + 1), and (*p*_1_, …, *p*_4_) are tuning parameters. The DRAM algorithm was used to get samples from the posterior distribution of equation-7 parameters, using data from the 15 days prior to the date from which it is desired to extrapolate. For a day *x* after the last one for which the data are available, the reported number of cumulative cases can be estimated as *C*_*R*_(*x*) = *C*(*x*)*/F* (*x*).

## 3 Results

Prior distributions and results of the model fits to the five data sets are summarized in Tables 1-15. The lowest Bayesian-information-criterion (BIC) value for the first wave in Santiago de Cuba was obtained with the model GRM+U (Table 2). The other lowest BIC values were obtained with: the model GRM+U for the second wave in Santiago de Cuba (Table 5), the model Gompertz+U for the first wave in Cuba (Table 7), the model SIR+U for the second wave in Cuba (Table 12), and the model Gompertz +U for the first wave in Spain (Table 13). In all the trials carried out, the BIC values obtained including the new model (+U) were lower than those calculated without including the new model.

We selected six results to provide visual evidence of what can be achieved with the new model. The Gompertz+U model visually follows the variability of the first wave data in Santiago de Cuba better than the Gompertz model (Figure 1). In figure 1B, the green-triangle line is remarkably close to the line of asterisks that represents the data. This green-triangle line corresponds to the maximum a posteriori (MAP) of the posterior density obtained by fitting the Gompertz+U model to the data of Santiago de Cuba (first wave). Figure 1 includes mean trajectory (blue) and the 95% credible intervals of drawn-from-posterior trajectories (light blue). The 95% credible intervals of the total estimated cases are represented in light red (Figure 1A). Fitting the Gompertz model to the same data shows a clearer difference between the fit and the data (Figure 1C).

**Figure 1:**
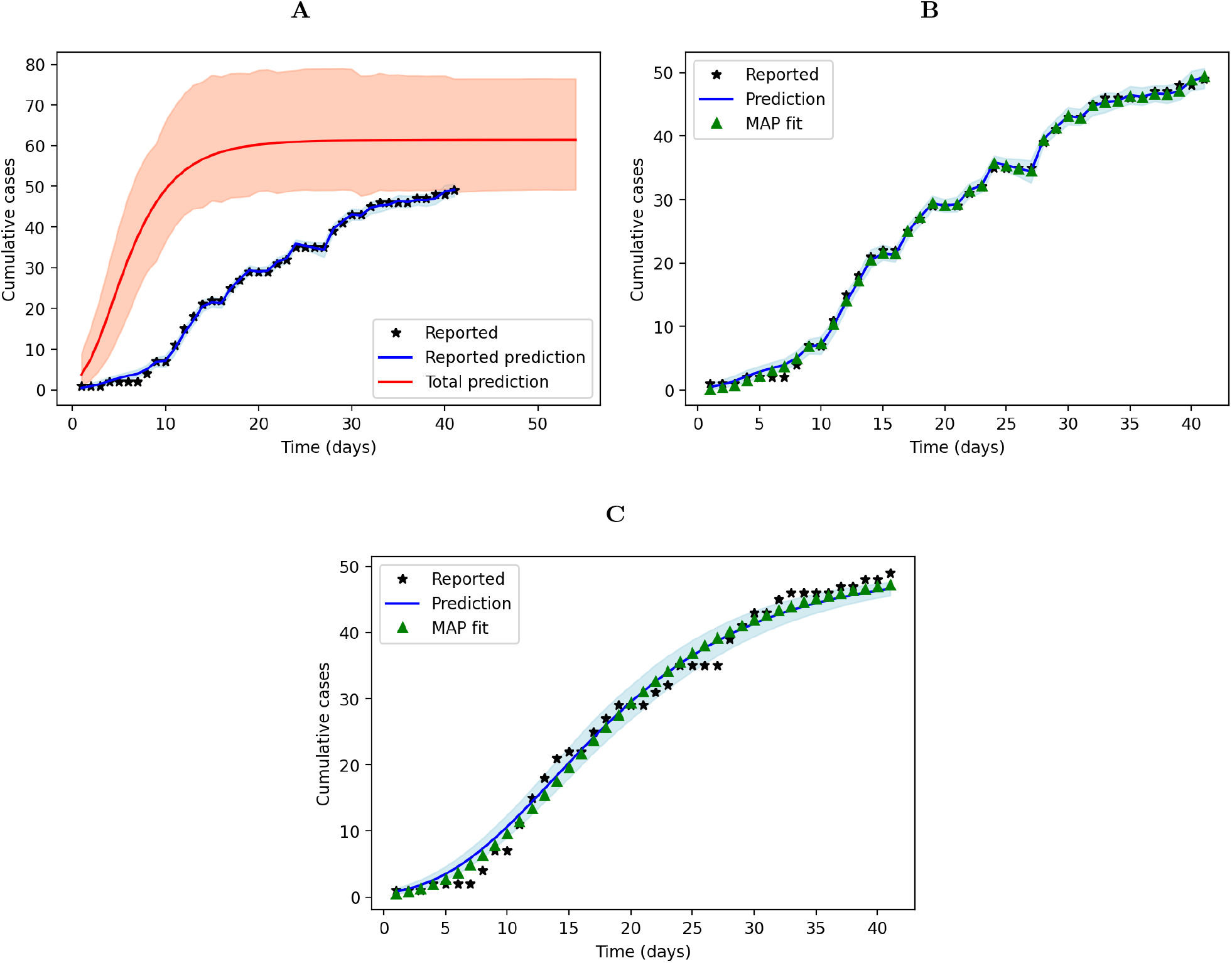
Gompertz-based models fitted to the data of the first epidemic wave in Santiago de Cuba. It includes the representation of the reported official numbers, the Gompertz+U fit (A), the Gompertz+U fit showing only the reported cases and highlighting the MAP fit (B), and the Gompertz fit to the data, highlighting the MAP fit (C). Shaded areas show the 95% credible intervals of posterior trajectories.

The first 75 of the 85 days analyzed in the second wave of Santiago de Cuba were taken to fit the models, and the remaining ten days were used to test model predictive ability. This division was carried out taking into account a change in the trend with a greater growth of new cases in the last ten days. The extrapolated Gompertz+U model was able to roughly follow this variation (Figure 2A). The extrapolation was carried out with equation 7 fitted to the estimated ratio of total to reported cumulative cases from 60 to 75 days (Figure 2B). The extrapolation of the Gompertz model was not able to follow the new trend (Figure 2C). Equation 7 was also fitted directly to the data from 60 to 75 days, without using another extra model, to extrapolate (Figure 2D).

**Figure 2:**
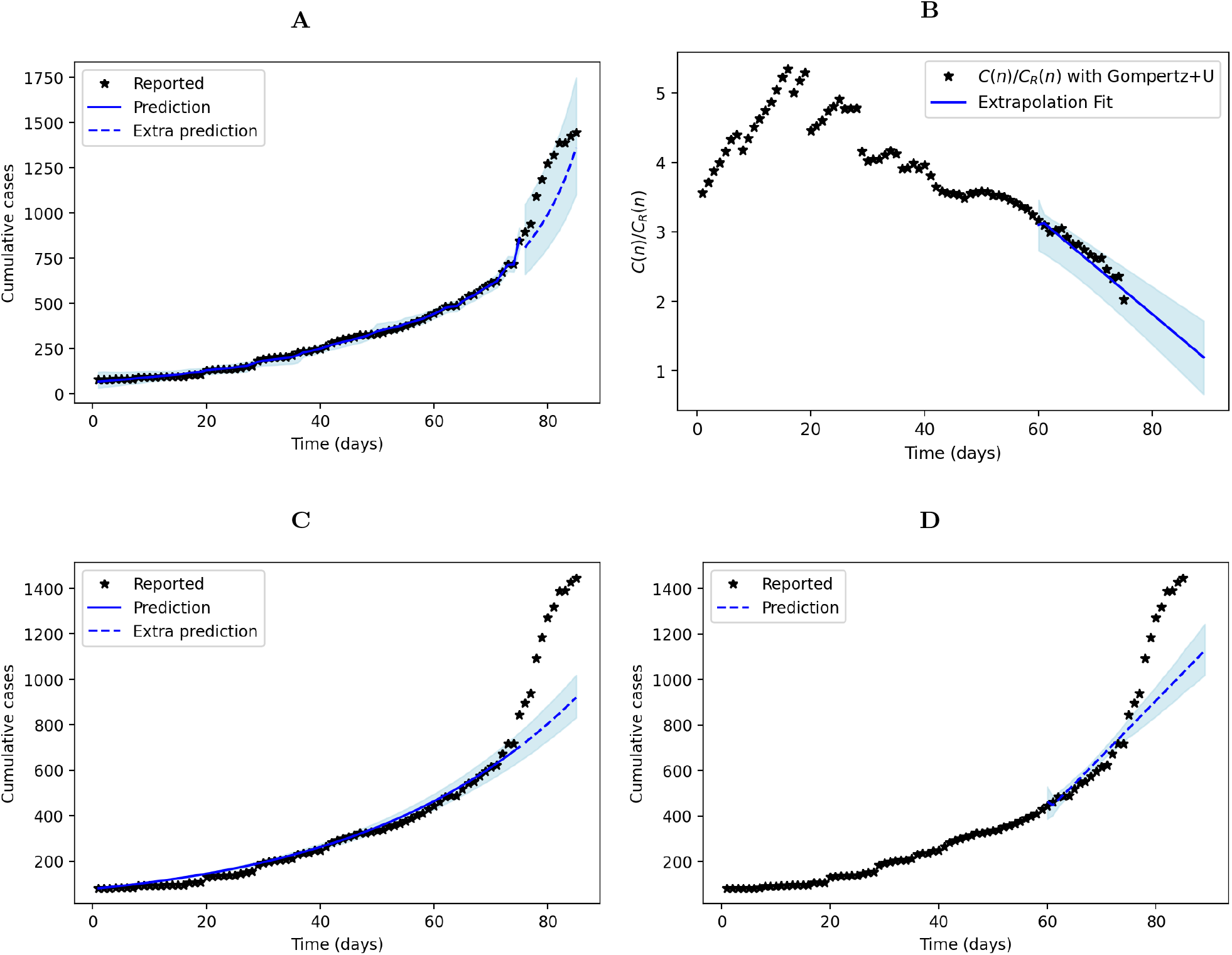
Gompertz-based models fitted to the data of the first 85 days of the second epidemic wave of Covid-19 in Santiago de Cuba, with extrapolation. The data is divided into 75 days to fit and ten days to extrapolate. Includes the fit of Gompertz+U model with extrapolation (A), the fit of the extrapolation function (equation 7) to total-cumulative-cases/reported-cumulative-cases ratio obtained with Gompertz+U model (B), the fit of Gompertz model with extrapolation (C), and the fit of the extrapolation function (equation 7) to the data directly (D).

## 4 Discussion

For all data sets studied, the best BIC values were obtained with some combination that includes the new model (Tables 1-15). This implies that the inclusion of the three new parameters *a, b* and *c*, does not overfit the resulting model, but is justified by the variability of the data. This result is promising if one takes into account that, for example, compared to the Akaike infomation criterion (AIC), the BIC tends to favor simpler models [33]. In most cases, the evidence in favor of the new model (+U) with respect to the reference model is classified as strong or very strong.

The proposed model is able to provide reasonable estimation of the number of unreported infected cases. The total number of accumulated cases on the 50th day of the first wave in Santiago de Cuba was previously estimated at 73 [8]. The Gompertz+U model estimates it at 60.3758 (95%*CI* : 51.5345 − 70.2727) (the estimate with all the daily credible intervals of the Gompert+U model with the data of the first wave in Santiago de Cuba can be visualized in Figure 1), the GRM+U model estimates it at 58.6924 (95%*CI* : 50.4297 − 63.5315) and the SIR+U model estimates it at 60.3758 (95%*CI* : 51.5345 − 70.2727). The approach of the present work has the advantage that it was not necessary to divide the wave into three parts for its analysis as in [8].

A previous report that considers the under-reporting [35] estimated for March 23, 2020, a total of 1 421 505 cases in Spain. For the same date, the Gompertz+U model estimates, for total cases, a 95% credible interval of 82 840 − 2 328 569 (it includes the estimated value in [35]), the GRM+U model estimates a 95% credible interval of 1 335 661 − 18 701 982 (it includes the estimated value in [35]), and the SIR+U model gives a 95% credible interval of 133 320 − 1 343 510 (5% below the estimated value in [35]). It is inferred that the choice of the deterministic model influences the quality of the estimation of the undetected cases.

The data for Spain are those with the highest number of reported cases among the five studied. Noise due to variability in reports is expected to have less influence on model-based results, compared to small-number data. In most cases, the absolute values of the parameters *b* and *c* were small compared to those of the other data (compare Tables 13-15 with Tables 1-12). For example, the absolute value of *b* for the Gompertz+U model in the first wave of Santiago de Cuba is more than 3000 times greater than the absolute value of *c* for the same model fitted to the data from Spain. This shows that the new model works properly with small variability as well as with large variability in reports.

The combinations obtained with the new model can predict the evolution of an ongoing epidemic wave, if a suitable extrapolation method is used. The example shown of successful extrapolation (Figure 2A) corroborates that the good fit of the new model to the data does not mean that it has a bad generalizability. Figure 2 illustrates a problem that is possible in practice. If a decision-maker asks the researcher to predict as accurately as possible the new cases in the next ten days, starting on the 75^th^ day of the second wave in Santiago de Cuba, in order to ensure the hospital capacities and all the necessary resources, it would be an underestimation if the Gompertz model (Figure 2C) or the extrapolation method applied directly to the data (Figure 2D) is used. In this case, only the extrapolation carried out from the Gompertz+U model offers an acceptable estimate. Nevertheless, the extrapolation function (equation 7) did not perform with the same quality in all trials. This suggests that it is necessary to observe the time-dependent dynamics of the actual-to-reported-cases ratio in order to choose the appropriate extrapolation function.

Based on the experience that the evolution of the first wave in Santiago de Cuba was favourable compared to the second one, we argue that the parameter *b* may be a quality criterion of the epidemiological research carried out in the region/country (the values of *b* in tables 1-3 are, at least, more than 25 times greater than the values of *b* in tables 4-6).

The new model can be combined with any deterministic model, making it possible to use it with SIRD (susceptible-infectious-recovered-dead), SEIR (susceptible-exposed-infectious-removed) and other compartmentalized variants. The forecast limitations are directly related to the limitations of the chosen deterministic model. In our case, the deterministic models Gompertz and GRM allow predicting the end of a wave, but they do not predict peaks of active infections or epidemic rebounds. Nevertheless, the Gompertz+U model shows good results in making short-term predictions.

In some trajectories drawn from posterior distributions of the new-model parameters, a growth of reported cases was observed towards the final days when the deterministic model already showed a plateau shape. This was not a reason to discard these regions in the analysis because we consider this phenomenon to be plausible, because sometimes samples are accumulated without being tested by real-time polymerase chain reaction, due to laboratory availability or priority of the authorities of one region over another.

Some current limitations of the new model are:

1. Active cases numbers are needed. For this reason it cannot be applied to the data provided by the World Health Organization in https://covid19.who.int without additional information.
2. It is assumed that the deterministic model captures the dynamics of the total number of cases in the region/country.
3. Several posterior peaks (multimodal posteriors) can be found in the estimation of the parameters.

The influence of the third limitation is reduced in this work by imposing limits on initial conditions of the model. Despite the above, we consider that the new model is a powerful tool to complement the information that decision-makers need to establish the appropriate policies to contain Covid-19.

We propose the following steps to generalize short-term predictions with the new model to data from an ongoing epidemic wave:

1. Collect the data of active and cumulative cases of the region/country where the epidemic wave is ongoing.
2. Fit the Gompert+U, GRM+U and SIR+U models to the collected data. Although in this article the Bayesian approach has been used for its advantages, it is possible to perform the fitting also with the maximum likelihood estimation or least-squares approaches.
3. Obtain the model-estimated ratio between the total and reported cumulative cases (*F*(*n*) = *C*(*n*)=*C_R_*(*n*)).
4. Choose a number of days (*D*(−) = 15 in this paper) to take the last reported data and extrapolate.
5. Extrapolate *F*(*n*) a number of days (*D*(+) = 15 in this paper) by fitting the equation 7 to the data of the days *D*(−).
6. Repeat steps 4 and 5, changing values until a visually adequate fit is obtained (see figure 2B).
7. Predict the number of cumulative cases with *C_R_*(*x*) = *C*(*x*)/*F*(*x*) where *x* is the day in *D*(+) for which the prediction is desired.

## 5 Conclusions

We propose a new mathematical model to describe the dynamics of the number of undetected cases for each reported case of SARS-CoV-2 infection in a region or country. The new model needs to be combined with a deterministic model that describes the evolution of the total number of cases in the selected region. In this study, we combined the new model with Gompertz growth, generalized logistic growth and SIR model, we fitted them to data from Cuba and Spain, and the results were better than the Gompertz, logistic and SIR models directly fitted to data. The proposed model deals well with variability in reports, estimates well the number of unreported infected and allows reasonable predictions. Although additional studies are needed to fully understand its parameters, the new model is already a useful tool for decision-makers to face waves of Covid-19.

## Data Availability

All data used are available.

https://covid19cubadata.github.io

https://cnecovid.isciii.es/covid19

